# Seasonality of non-SARS, non-MERS Corona viruses and the impact of meteorological factors

**DOI:** 10.1101/2020.07.15.20154146

**Authors:** Olympia E. Anastasiou, Anika Hüsing, Johannes Korth, Fotis Theodoropoulos, Christian Taube, Karl-Heinz Jöckel, Andreas Stang, Ulf Dittmer

**Affiliations:** Institute for Virology, University Hospital Essen, University of Duisburg-Essen, Germany; Institute of Medical Informatics, Biometry and Epidemiology, University Hospital Essen, University Duisburg-Essen, Essen, Germany; Department of Nephrology, University Hospital Essen, University of Duisburg-Essen, Germany; Department of Pulmonary Medicine, University Hospital of Essen - Ruhrlandklinik, Essen, Germany; Centre for Clinical Studies (ZKSE), Institute for Medical Informatics, Biometry and Epidemiology, Medical Faculty, University Duisburg-Essen, Essen, Germany

## Abstract

**Background:** Seasonality is a characteristic of some respiratory viruses. The aim of our study was to evaluate the seasonality and the potential effects of different meteorological factors on the detection rate of the non-SARS Corona Virus detection by PCR.

**Methods:** We performed a retrospective analysis of 12763 respiratory tract sample results (288 positive and 12475 negative) for non-SARS, non-MERS Corona viruses (NL63, 229E, OC43, HKU1). The effect of seven single weather factors on the Corona virus detection rate was fitted in a logistic regression model with and without adjusting for other weather factors.

**Results:** Corona virus infections followed a seasonal pattern peaking from December to March and plunging from July to September. The seasonal effect was less pronounced in immunosuppressed patients compared to immunocompetent. Different automatic variable selection processes agreed to select the predictors temperature, relative humidity, cloud cover and precipitation as remaining predictors in the multivariable logistic regression model including all weather factors, with low ambient temperature, low relative humidity, high cloud cover and high precipitation being linked to increased Corona virus detection rates.

**Conclusions:** Corona virus infections followed a seasonal pattern, which was more pronounced in immunocompetent patients compared to immunosuppressed. Several meteorological factors were associated with the Corona virus detection rate. However, when mutually adjusting for all weather factors, only temperature, relative humidity, precipitation and cloud cover contributed independently to predicting the Corona virus detection rate.

## Introduction

Respiratory tract infections (RTI) are an important contributor to overall morbidity and mortality, with lower RTI being the fourth most frequent cause of death worldwide (1). Respiratory viruses are also an important cause of outbreaks and epidemics, including the widespread and in-depth studied Influenza virus (2) but also the recently isolated Corona virus, SARS-CoV-2, which continues to have a devastating effect on public health and global economy (3, 4).

Seasonality is a prominent characteristic of many viral RTI. In temperate climate regions viral RTI reach their peak in winter and in tropical areas during the rainy season (5-8). It has been postulated that the underlying causes of their seasonality are both virus- and host-related. Meteorological factors can influence virus survival but also modulate human behavior (5, 9) and host immune responses (10). Host-related factors can influence not only the incidence but also the clinical course of an infection. Previous studies indicate that immunosuppression is a risk factor for an unfavorable outcome in patients with respiratory infections (11-13). Furthermore, there is evidence of prolonged virus shedding in immunocompromised hosts (14), the clinical and epidemiological importance of which remains unclear.

Interestingly, meteorological factors seem to play a part in the seasonal distribution of some respiratory viral pathogens (6, 7). Preprints of recent studies suggest a potential association between SARS-CoV-2 incidence and climate conditions (15-17) but the evidence remains inconclusive and the association is not supported by all available data (18). In addition, seasonality and climate dependence of SARS-CoV-2 could not be adequately studied until now because the ongoing pandemic lasts only a couple of months and was dramatically influenced by contact distancing measures. In-depth analysis of the seasonal pattern of non-SARS, non-MERS Corona viruses in conjunction with meteorological factors might therefore provide important insights into the biology of Corona viruses in general.

The aim of the present study was to evaluate the seasonality and the potential effect of different meteorological factors on the detection rate of the non-SARS, non-MERS Corona viruses. Furthermore, we focused on potential differences in the seasonality of Corona viruses in immunocompetent vs immunosuppressed hosts.

## Materials and Methods

We performed a retrospective analysis of 12763 samples (288 positive and 12475 negative), including nose/throat swabs, tracheal aspirates, bronchoalveolar lavages, tested in the Institute for Virology of University Hospital Essen, Germany from June 2013 to December 2019. The samples were tested with the respiratory viral panel (FTD, Siemens, Erlangen, Germany) according to the manufacturer’s instructions, for the detection of non-SARS, non-MERS Corona virus (NL63, 229E, OC43, HKU1). The analysis included all tested samples, meaning that a patient could contribute more than once over time, with the following exception: repeated positive samples from the same individual collected within two weeks of each other were removed. The term “Corona virus” corresponds to non-SARS, non-MERS Corona viruses in our manuscript, unless specifically otherwise noted (e.g. SARS-CoV-2). Nucleic acid extraction was performed using MagNA pure (Roche, Mannheim, Germany). Demographic and clinical data were taken from patient charts. A quarter of our cases (n=3254, 26.1%) and a third of our positive cases (n=92, 31.9%) were generated by immunosuppressed individuals (patients with hematological or oncological malignancies under chemotherapy, solid organ transplant recipients, patients after allogeneic human stem cell transplantation).

Meteorological data were obtained for each day of the study period from the weather station of Essen Bredeney, Germany through the server of “Deutscher Wetterdienst”. The data included daily average temperature, daily average relative humidity, precipitation, daily average wind speed, sunlight hours, daily average cloud cover and daily average atmospheric pressure. We expressed the association between continuous weather variables and the corona virus detection rates by increments of 5 units on the corresponding scales of the variables if not otherwise specified. The meteorological factors in Essen, Germany for the duration of the study are presented in Supplement (Figure S1). This retrospective study was carried out in accordance with the Declaration of Helsinki and the guidelines of the International Conference for Harmonization for Good Clinical Practice.

### Data analysis

Virus detection rate was modelled via logistic regression with a seasonal effect and/or weather factors as explanatory variables. Seasonal variation in virus detection rate was fitted for each year separately with a cosinus function with 1-year frequency length together with a sinus curve, and all variables with a p-value below 0.157 [i.e. improving the model quality according to the Akaike-information criterion (AIC)] were combined into a single year-specific seasonal score. The formula can be found in the supplement (Supplement Table S1).

Weather was represented as daily average values locally assessed 10 days prior to the virus measurement to account for incubation time (19-21) and time from the first symptoms to Corona virus diagnostics (10 days lag-time). Weather measurements and fitted seasonal effect were compared via Pearson’s correlation analysis.

The effect of single weather factors on the Corona virus detection rate was fitted in a logistic regression model with and without adjusting for other weather factors and seasonality. Variable selection was applied to the weather factors with and without adjustment for season as backward, forward, and stepwise selection, targeting statistical model optimization as improvement of AIC. Selected effects were also estimated in patients stratified according to their immune status (immunocompetent vs immunosuppressed).

The monthly Corona virus detection rate as the number of positive samples per tested samples per month was compared with the corresponding sum of predicted values from the logistic model. Additionally the Corona virus detection rate was compared with the sum of predicted values within deciles of the predicted values in a calibration plot. We are calculating and reporting confidence intervals to assess the precision of our estimates because our goal is estimation and not significance testing. We wish to avoid publication bias by preferential reporting of significant results. Instead, we judge the value of our estimates by their precision and validity (22, 23). All statistical analyses were performed using SAS v.9.4 (SAS Institute, Cary, North Carolina, USA).

## Results

### Corona virus infections followed a seasonal pattern

Since 2013 we generated 12763 test results for the detection of the non-SARS, non-MERS Corona viruses (NL63, 229E, OC43, HKU1) by multiplex PCR technology. Corona virus infections followed a clear seasonal pattern among our patients. Although Corona viruses were detected throughout the years, detection rates were at their peak from December to March and at their nadir from July to September. In October and November, the number of positive cases was low in most years, usually lower than in May. As shown in Figure 1 the monthly detection rate differed from year to year. The Corona virus detection rate could be predicted well with a mathematical model including the seasonal effect alone, as well as a model only including seven weather factors (Figure 1A and 1B respectively). The model based on seasonal effect was slightly superior in this aspect, fitting the data more accurately with concordance-statistics c= 0.74 for model A and c=0.68 for model B and as shown in calibration plots (Figure S2) in the supplement.

**Figure 1:**
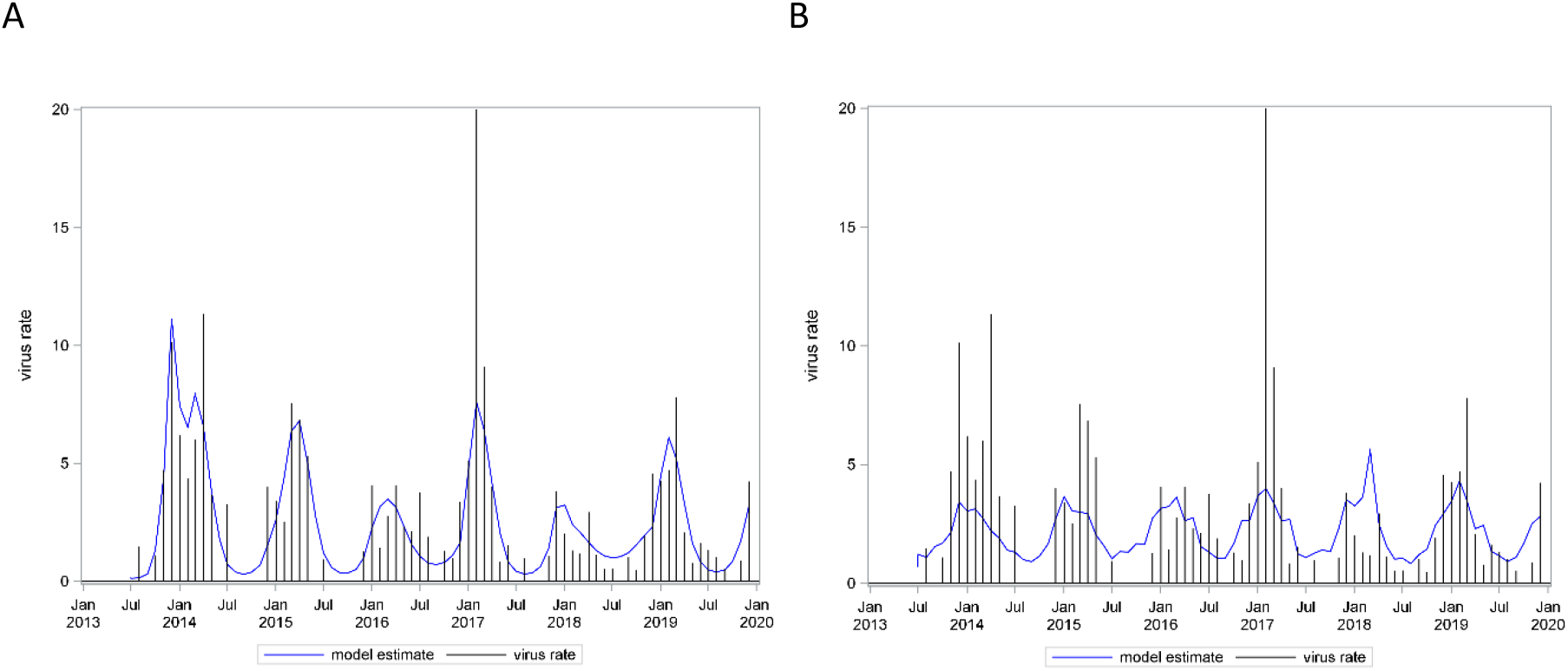
Corona virus infections followed a seasonal pattern. The needles in A and B panels show the monthly detection rate. The curve in panels A and B depict the models describing a seasonality effect (A) and a combined effect of seven weather factors (daily average ambient temperature, relative humidity, wind speed, cloud cover, atmospheric pressure, precipitation and number of sunlight hours) (B) on Corona virus detection rate, respectively. Both models had an adequate fit, however a better model fit was obtained using seasonality to predict virus detection rates.

### Low ambient temperature, minimum sunlight hours per day, high relative humidity, wind speed, cloud cover and precipitation were each associated with higher detection rates of Corona virus

We next analyzed the effect of weather factors on Corona virus detection rates. Since Corona viruses including MERS and SARS have an incubation time between 2-11 days (19-21), we associated the weather factors with infection rates that were diagnosed 10 days later to account not only for the incubation time but also the time passing between the first symptoms and implementation of diagnostics. The crude effect of single weather factors on the Corona virus detection rate showed associations of lower ambient temperature [Odds Ratio (OR) 0.68, 95% confidence interval (CI) 0.62-0.74], minimum sunlight hours per day (OR 0.93, 95% CI 0.9-0.96), higher relative humidity (OR 1.04, 95% CI 1-1.09) and wind speed (OR 1.14, 95% CI 1.04-1.24), cloud cover (OR 1.12, 95% CI 1.05-1.19) and precipitation (OR 1.14, 95% CI 1.03-1.27) with higher detection rates of Corona virus. Higher atmospheric pressure showed only a weak decreasing effect (Figure 2). We expressed the association between continuous weather variables and the corona virus detection rates by increments of 5 units (instead of one unit) on the corresponding scales of the variables daily average temperature, daily average relative humidity, daily average atmospheric pressure and daily precipitation.Since weather factors are correlated with each other to a greater or lesser degree (from correlation (ρ) of 0.01 between temperature and rain to ρ= 0.80 between sunlight and cloud cover – see Table S2 in the supplement), we performed the same analysis for each weather factor after adjusting for all the others. After adjustment, the above-mentioned effects were preserved for ambient temperature (OR 0.62, 95% CI 0.55-0.69), cloud cover (OR 1.13, 95% CI 1.02-1.26) and precipitation (OR 1.25, 95% CI 1.1-1.41). However, in this adjusted analysis, wind speed and hours of sunlight lost impact, low relative humidity (OR 0.84, 95% CI 0.79-0.91) was now associated with higher virus rates, and the atmospheric pressure showed a weak increasing effect.

**Figure 2:**
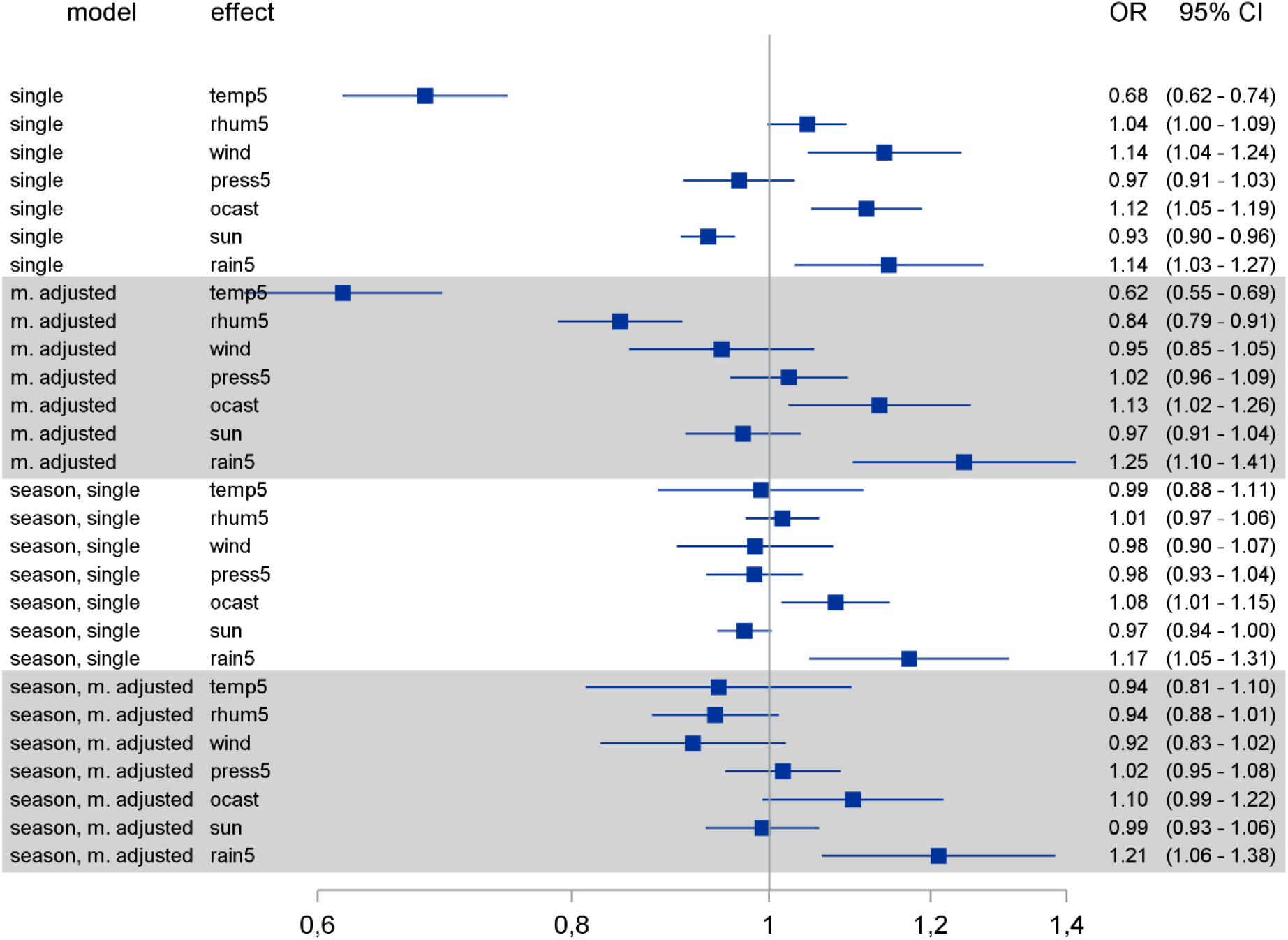
Logistic regression models were calculated for each weather factors with and without adjusting for other weather factors and/or seasonality. OR: Odds Ratio, CI: confidence interval. Temp5: daily average temperature [per 5°C change], rhum5: daily average relative humidity [per 5% change], wind: daily average wind speed [per 1m/s change], press5: daily average atmospheric pressure [per 5hPa change], ocast: daily average cloud cover, sun: sunlight hours daily, rain5: precipitation daily [per 5mm change].

Most weather factors demonstrate a seasonal pattern (Figure S1). To account for the seasonality effect, we performed the same analysis for each weather factor after adjusting for seasonality and additionally after adjusting for seasonality and the other weather factors. After adjusting for seasonality all associations were maintained as before, but largely lost impact. Only the effects of cloud cover and precipitation remained robust. After adjusting for both seasonality and all other weather factors, the point estimates followed the same pattern as in the multiple adjustment above, with cloud cover and precipitation clearly showing increasing effects on virus detection rates (Figure 2). The fact that ambient temperature lost its impact on the viral detection rate in the analysis adjusted for seasonality might be explained by the strong association between temperature and season (Supplement Figure S1 and Table S2).

### Temperature, relative humidity, precipitation and cloud cover were independently associated with the Corona virus detection rate

Next, we aimed to calculate a reduced predictive model for the Corona virus detection rate based on the weather factors. Different automatic variable selection processes agreed to select the predictors ambient temperature, relative humidity, cloud cover and precipitation as remaining predictors in the logistic regression model (Figure 3). All above-mentioned factors were independently associated with the Corona virus detection rate, with low ambient temperature, low relative humidity, high cloud cover and high precipitation being linked to increased Corona virus detection rates.

**Figure 3:**
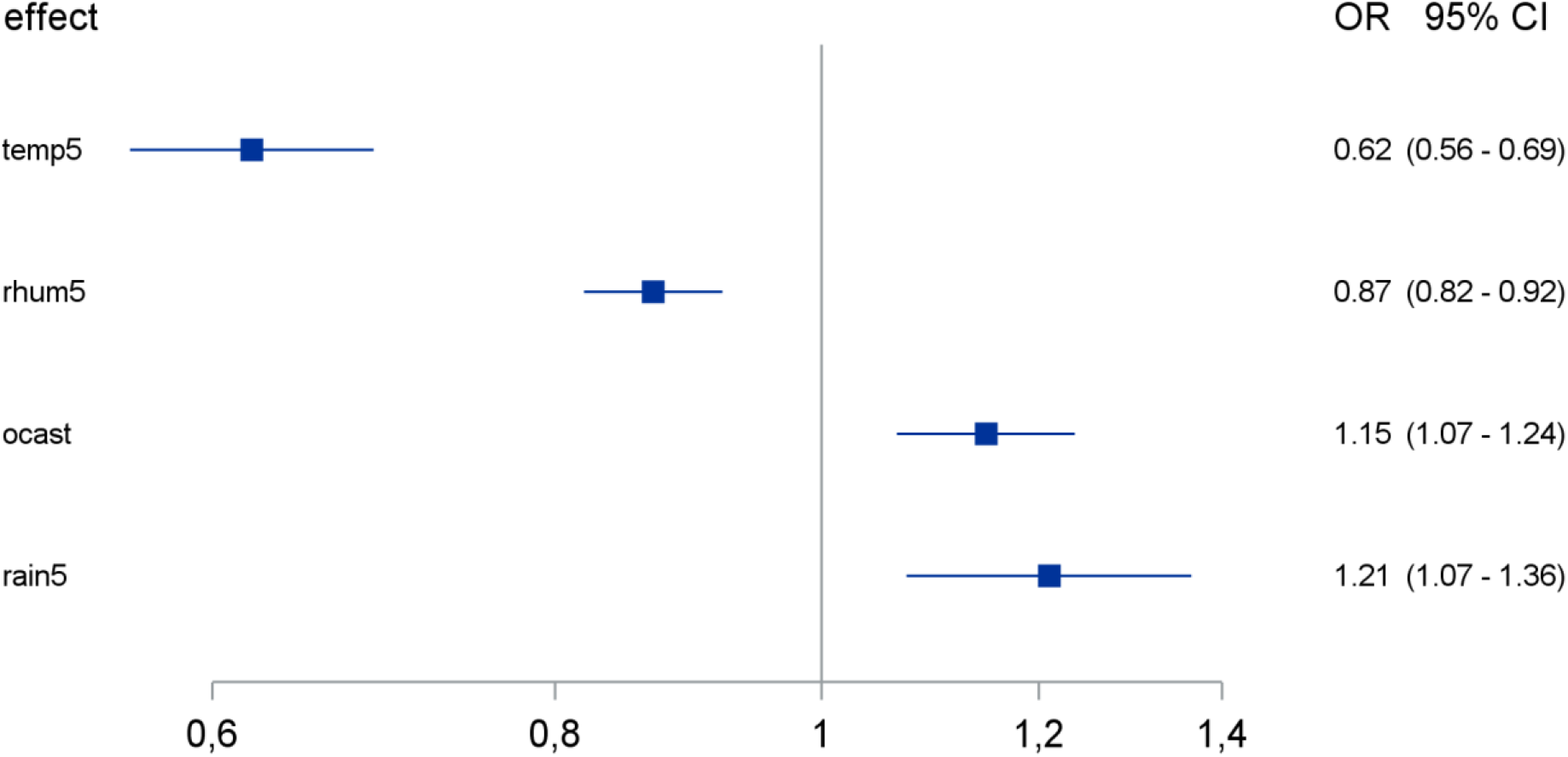
Effect of weather factors on Corona virus detection rate according to a reduced logistic regression model of the combined effect of weather factors. OR: odds ratio, CI: confidence interval. Temp5: daily average temperature [per 5°C change], rhum5: daily average relative humidity [per 5% change], ocast: daily average cloud cover, rain5: precipitation daily [per 5mm change].

### The seasonality of Corona virus infections was less pronounced in immunosuppressed patients

A quarter (26.1%) of our patients consisted of immunosuppressed individuals generating a third (31.9%) of our positive cases. The group of immunosuppressed patients included patients with hematological or oncological malignancies under chemotherapy, solid organ transplant recipients and patients after allogeneic human stem cell transplantation. To evaluate the potential impact of immunosuppression on the seasonality of Corona virus detection, we divided our patients in immunosuppressed and immunocompetent patients and used logistic regression models to calculate the seasonal effect and the effect of weather factors through multivariable logistic regression model (Figure 4) selected weather factors on the Corona virus detection rates in both groups. As demonstrated in Figure 4, the seasonality effect was less pronounced in cases involving immunosuppressed patients compared to their immunocompetent counterparts, but the selected weather factors had comparable effects in both groups.

**Figure 4:**
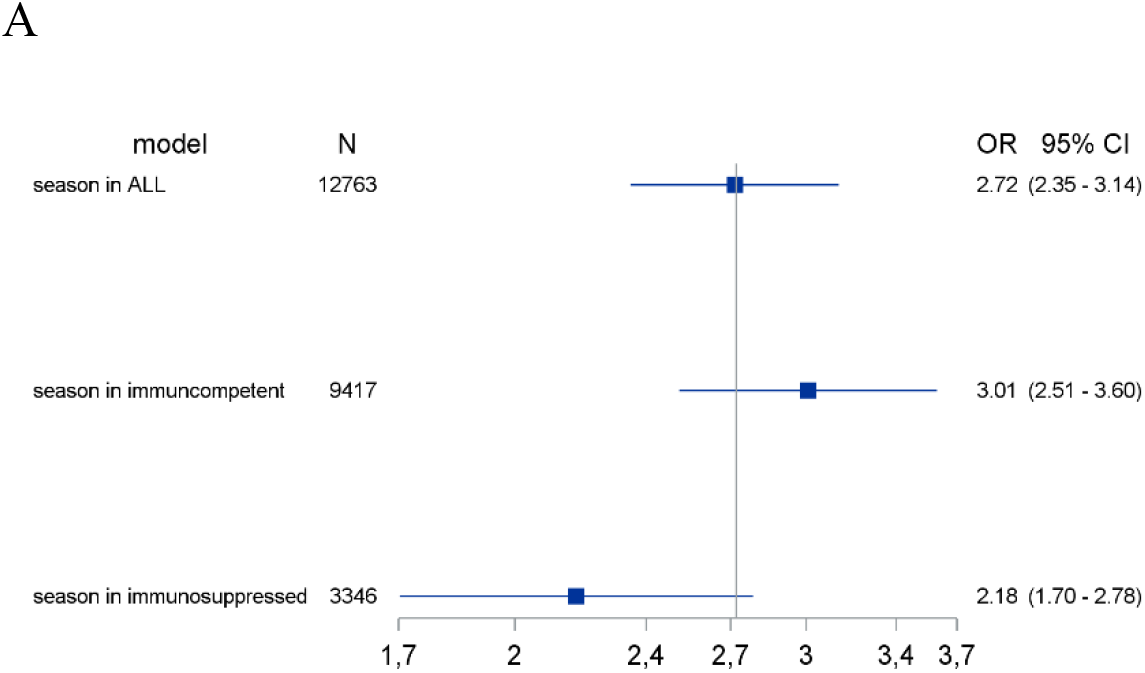
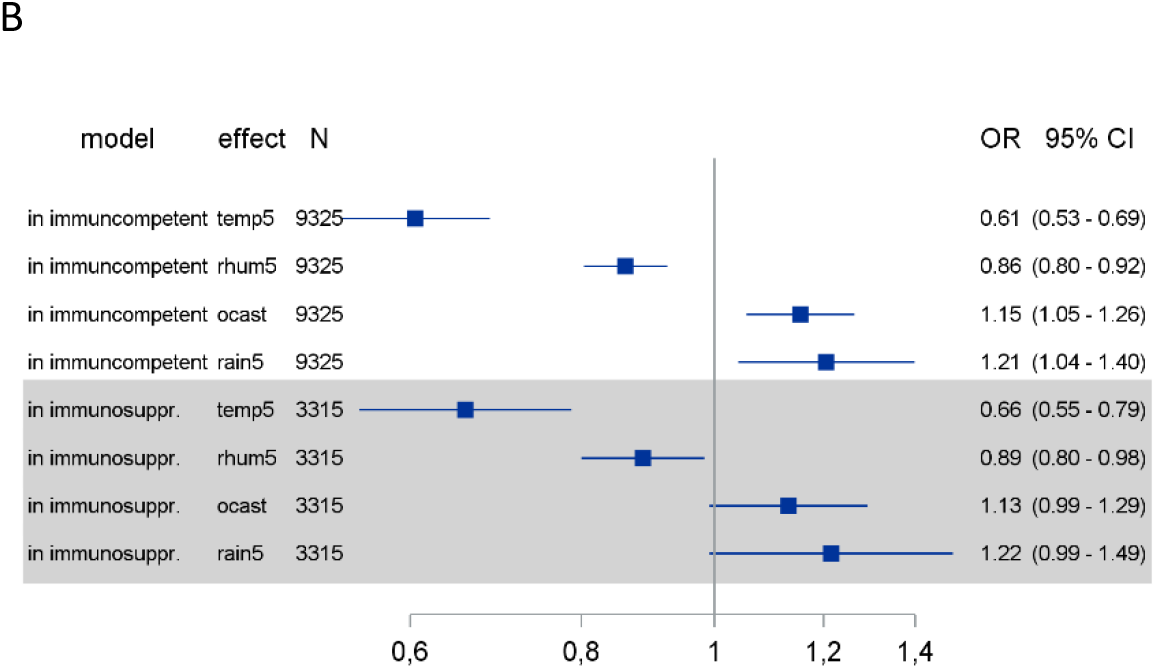
The seasonality and weather influence of Corona virus detection rates was compared between immunosuppressed and immunocompetent patients. Logistic regression models were calculated for the effect of seasonality (A) and the effect of selected weather factors (B) on the Corona virus detection rate in immunosuppressed and immunocompetent patients. OR: odds ratio, CI: confidence interval. Temp5: daily average temperature [per 5°C change], rhum5: daily average relative humidity [per 5% change], ocast: daily average cloud cover, rain5: precipitation daily [per 5mm change] season: grand average year-specific seasonal effect.

### “Off-season” Corona virus detection was more frequent in immunosuppressed patients

The Corona virus detection rate demonstrated a pronounced seasonal pattern, but even in the time period with the lowest detection rate (summer to fall), we could detect some Corona virus positive cases. Thus, we aimed to characterize these cases from a clinical point-of-view.

As shown in Figure 1, the frequency of Corona virus detection was at its nadir from July to September. When analyzing the clinical characteristics of patients with Corona virus infection during its nadir (n=33), we observed that the majority of positive cases (n=22, 66%) were immunosuppressed. This included eleven solid organ transplant recipients, six patients after allogeneic bone marrow transplantation and five patients on chemotherapy due to malignant diseases. Information on the travel history of these patients in the last two weeks before Corona virus detection were available for 29 cases and all but one patients had not been abroad. Thus, they must have acquired their Corona virus infection in their local environment, which might have happened weeks before the positive test result.

Among our patients, data on viral persistence were limited, since follow-up was not consistently performed in most cases. In six cases, we observed viral persistence lasting for more than a month (from 34 to 116 days). Five out of theses 6 patients were immunosuppressed.

## Discussion

Corona virus infections with the viruses NL63, 229E, OC43 and HKU1 followed a seasonal pattern among our patients. Although Corona viruses were in principle detected throughout the year, its detection rate was at its peak from December to March and at its nadir from July to September. “Off-season” detection of Corona virus was more frequent in immunosuppressed patients. Low ambient temperature, few sunlight hours per day, high relative humidity, wind speed, cloud cover and precipitation were each individually associated with high detection rates of Corona viruses. In a multivariable model including all weather factors temperature, relative humidity, precipitation and cloud cover were independently associated with the Corona virus detection rate

Seasonality is commonly observed in respiratory infections. Its form and impact depends on the individual pathogen and the climate zone under evaluation (5-8). Our study focused on the seasonal non-SARS, non-MERS Corona virus detection rate in Germany, a temperate climate country. We observed a distinctive seasonal pattern, peaking in winter and plunging in the summer. Our data on seasonality are largely consistent with previous studies evaluating the seasonality of Corona virus (24, 25). However, we show for the first time that data sets on detections rates over many years fit perfectly with a mathematical model of seasonality and weather dependence.

Seasonality is obviously a characteristic of Corona virus infections. Among our patients, its detection rate was at its peak from December to March and at its nadir from July to September. This pattern has similarities but also differences compared to Influenza virus detection. Both viruses peak in the winter, but Influenza virus has a markedly narrow time interval of detection. It is highly prevalent from January to March, but disappears in late spring (usually April), summer and autumn (apart from very few isolated cases) as shown in previous studies (6, 7) and in data from the German disease control and prevention agency (Robert-Koch-Institute) (Supplement, Figure S3). Corona virus on the other hand lingers longer during the spring months (until May/June) and appears earlier in the autumn (in November), without completely disappearing over the summer.

A seasonal effect on Corona virus detection rates was present in both immunocompetent and immunosuppressed patents, but it was less pronounced in the immunosuppressed group. “Off season” detection of Corona virus was more frequent in immunosuppressed patients. Focusing on the “off-season” Corona virus cases, we found that 66% of them were observed in immunosuppressed individuals, although immunosuppressed patients accounted for only 26% of our cases. We have only limited data on the persistence of Corona viruses, but prolonged viral shedding was observed among our patients and it involved mainly immunosuppressed individuals. This is consistent with current knowledge about respiratory virus shedding in humans (14, 26) and has been reported in SARS-CoV-2 infection as well (27). Both phenomena are interesting from a clinical and epidemiological perspective. Corona virus infections can be detected in the summer months, albeit at very low infection rates. Infections were especially found in immunosuppressed patients, who were reported to be at risk for unfavorable outcomes in respiratory infections (11-13). In addition, prolonged viral shedding may favor the wider distribution of the virus and the emergence of viral variants as it has been reported for other viruses (28). Immunosuppressed patients may also serve as a reservoir for the virus in the summer. Thus, immunosuppressed hosts may sustain viral replication and spread under otherwise unfavorable external conditions for Corona viruses.

We also evaluated the association of meteorological conditions and Corona virus detection rates. Taken individually, low ambient temperature, minimum sunlight hours per day, high relative humidity, wind speed, cloud cover and precipitation were associated with higher detection rates of Corona viruses 10 days later. Similar weather effects, to a lesser or greater degree, have been described for other respiratory viruses, such as Influenza virus and respiratory syncytial virus (7). In a pediatric population study, the authors looked at the potential effects of some weather factors (temperature, wind velocity, relative humility) on the non-SARS, non-MERS Corona virus detection rate. Each variable was individually evaluated for its potential effect and indeed a negative correlation between temperature and the Corona virus detection rate and a positive one for wind velocity and relative humidity were observed (6). However, meteorological factors are not independent from one another. Using a multivariable logistic regression analysis and including seven weather factors, we found that only temperature, relative humidity, cloud cover and precipitation were associated with the viral detection rate. Interestingly, relative humidity seemed to have the opposite effect in a multivariable model compared to the univariate one; namely, low relative humidity was associated with higher viral detection rate. The observed effects of temperature and relative humidity are consistent with laboratory data for another Corona virus. SARS-CoV-1 had a better chance remaining infectious in a low temperature and low humidity environment (29). Our results demonstrate that the Corona virus detection rate is indeed associated with meteorological factors but also underline the potential pitfalls of analyzing the effect of single factors, without factoring other interdependent factors in a complex system.

Experimental data on the link between Corona virus transmission and weather factors are largely missing. However, experiments performed with another respiratory virus, e.g. Influenza viruses, provide some insight about the possible mechanistic explanation of these associations. We found a clear correlation of Corona virus detection rates and cloud cover or sun light hours. How can this be explained? Solar radiation, which is dependent on cloud cover and sunlight at a given location, has been shown to act virucidal against Influenza virus (30). The virucidal effect of ultraviolet radiation has also been recently demonstrated on SARS-CoV-1 (31). We also demonstrated a link between temperature and relative humidity and Corona virus detection rates. Temperature can influence virus stability through inactivation of proteins and nucleic acids (32) but may also influence the host’s defense systems. Cooling and drying of the nasal epithelium inhibits mucociliary clearance and viral phagocytosis (33, 34), thus facilitating infections. Low relative humidity can also promote Influenza infection and transmission. At lower relative humidity, salts within aerosols tend to crystallize out of the solution leading to higher virion stability (35), while respiratory tract droplets stay suspended longer and end in the lower respiratory tract more often, thus increasing both the risk of transmission and the risk for an unfavorable infection outcome (32).

High precipitation was associated with increased Corona virus detection rates among our patients, which seems counterintuitive, when one considers the association of lower relative humidity with higher Corona virus detection rates. Interestingly, the factors precipitation and relative humidity are not as closely linked (see Supplement, Table S2) as one would expect. A possible explanation is that precipitation does not directly affect viral infectivity or the host’s immune defenses, but mainly influences human behavior, e.g. through increased indoor congregation on a rainy day. Indeed, a similar effect has been described in a study focusing on Influenza, where a significant positive association was observed between extreme precipitation and emergency room visits for Influenza (36).

Both seasonality and the combined effect of weather factors are reliable predictors of Corona virus detection rates as shown in the models depicted in Figure 1. This is not surprising, since most weather factors demonstrate a pronounced seasonal pattern (Figure S1). Still, the model based on seasonal effects was slightly superior to the weather model in predicting the viral detection rate. Meteorological factors such as temperature can influence the virus itself, including viability and infectivity, but also the effect the host’s immune response (10, 37). Furthermore, they can also modulate human behavior increasing or decreasing the risk of transmission. Factors potentially contributing to this effect include but are not limited to indoor/outdoor activities, indoor heating, crowding and the impact of melatonin and vitamin D levels on the host’s immune defense (5, 9). The superiority of seasonality as a predictive factor for infection rates suggests that beside the significant correlation with several weather factors, also social and cultural norms that modify human behavior influence viral spread.

A limitation of our study was that we used a convenience sample, namely patients seeking treatment at our hospital. This leads to an exclusion of infected but asymptomatic individuals and probably to a marked underestimation of mildly symptomatic individuals, meaning that the true number of Corona virus-infected individuals in the general population at any given time is probably much higher. Furthermore, due to the nature of our study we cannot offer mechanistic explanations for the associations we observed. However, data on the association of weather factors and Corona virus detection rates are very limited, but very important to predict the ongoing SARS-CoV-2 pandemic. Our study describes, for the first time, the effect of the interplay of seasonality and several weather factors on the non-SARS, non-MERS Corona viruses detection rate and indicates that some but not all weather factors are independently associated with it, providing valuable insight in the seasonal pattern of Corona viruses in general. An association between meteorological factors and the SARS-CoV-2 detection rate has been suggested (15-17) but the evidence remains to date inconclusive (18). The epidemiological situation for SARS-CoV-2 is being further complicated due to drastic lockdown measures all over the world. Our analysis of the seasonal pattern of the non-SARS Corona viruses in conjunction with a potential association with meteorological factors might provide valuable information for the ongoing pandemic. Furthermore, data on the temporal pattern of Corona virus infections in immunosuppressed patients in the literature are limited. Our study allows a first direct comparison of the seasonal pattern and weather effects on the Corona virus detection rate in immunosuppressed versus immunocompetent patients.

In conclusion, Corona virus infections followed a clear seasonal pattern among our patients, which was more pronounced in immunocompetent patients compared to the immunosuppressed. Several meteorological factors were associated with the Corona virus detection rate. However, after multiple adjusting for all weather factors, only temperature, relative humidity, precipitation and cloud cover remained independently associated with the Corona virus detection rate.

## Data Availability

---------

## Supplement

**Figure S1:**
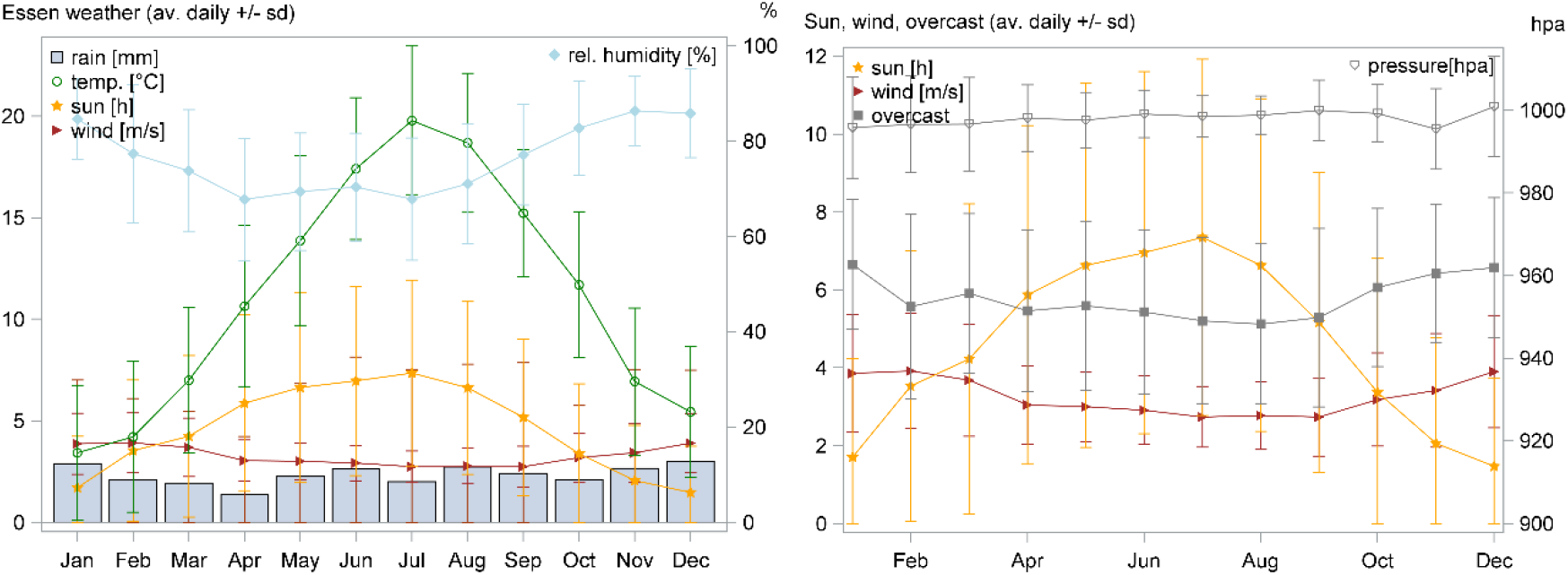
Weather factors in Essen, Germany. Figure S1A and B depict the average monthly weather factors in Essen, Germany from June 2013 to December 2019. Values are presented as mean±SD. SD: standard deviation.

**Table S1:** Estimated equation for the outcome season concerning the Corona virus detection rate.

| Year | Estimated equation for the outcome season |
| --- | --- |
| 2013 | $1.5789 * \cos((6.283185308/52)*(week - 9 - 0.5)) - 2.1464 * \sin((6.283185308/52)*(week - 9 - 0.5))$ |
| 2014 | $1.7072 * \cos((6.283185308/52)*(week - 9 - 0.5))$ |
| 2015 | $1.4852 * \cos((6.283185308/52)*(week - 9 - 0.5)) + 0.4968 * \sin((6.283185308/52)*(week - 9 - 0.5))$ |
| 2016 | $0.8244 * \cos((6.283185308/52)*(week - 9 - 0.5))$ |
| 2017 | $1.5616 * \cos((6.283185308/52)*(week - 9 - 0.5)) - 0.6142 * \sin((6.283185308/52)*(week - 9 - 0.5))$ |
| 2018 | $0.2456 * \cos((6.283185308/52)*(week - 9 - 0.5)) - 0.4102 * \sin((6.283185308/52)*(week - 9 - 0.5))$ |
| 2019 | $1.1880 * \cos((6.283185308/52)*(week - 9 - 0.5)) - 0.7662 * \sin((6.283185308/52)*(week - 9 - 0.5))$ |

**Figure S2:**
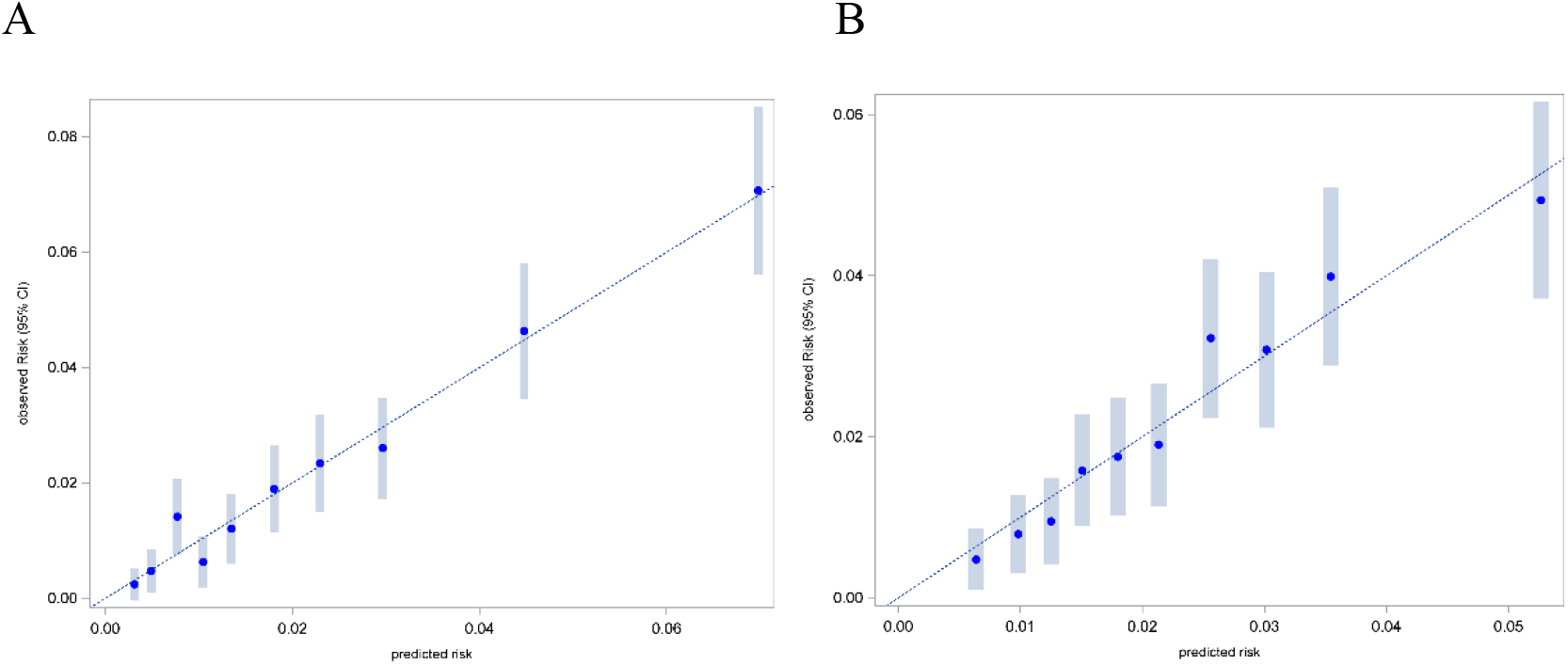
Panels A and B depict the calibration plot for the prediction of Corona virus detection rate in deciles of predicted values for a model based on a year-specific seasonality effect (A) and a model based on the combined effect of all weather factors (B) respectively.

**Table S2:** Correlation between weather factors.

| $\rho$<br>95% CI | Seasonal<br>effect | Temperature<br>(°C) | Wind<br>speed<br>(m/s) | Atmospheric<br>pressure<br>(hpa) | Relative<br>Humidity,<br>% | Precipitation,<br>mm | Sunlight<br>hours | Cloud<br>cover<br>(okta) |
| --- | --- | --- | --- | --- | --- | --- | --- | --- |
| Seasonal<br>effect | - | -0.62<br>(-0.65-<br>-0.59) | 0.25<br>(0.2-0.29) | -0.07<br>(-0.12--0.03) | 0.10<br>(0.06-0.15) | -0.03<br>(-0.07-0.02) | -0.21<br>(-0.26-<br>-0.17) | 0.1<br>(0.06-0.15) |
| Temperature<br>(°C) | -0.62<br>(-0.60-<br>-0.59) | - | -0.22<br>(-0.27-<br>-0.18) | 0.02<br>(-0.02-0.07) | -0.52<br>(-0.55-<br>-0.48) | -0.01<br>(-0.05-0.04) | 0.50<br>(0.47-0.54) | -0.25<br>(-0.29-<br>-0.2) |
| Wind speed<br>(m/s) | 0.25<br>(0.20 -<br>0.29) | -0.22<br>(-0.27-<br>-0.18) | - | -0.37<br>(-0.41--0.33) | 0.04<br>(-0.00-0.09) | 0.25<br>(0.21-0.29) | -0.23<br>(-0.27-<br>-0.18) | 0.18<br>(0.14-0.23) |
| Atmospheric<br>pressure<br>(hpa) | -0.07<br>(-0.12 -<br>-0.03) | 0.02<br>(-0.02-0.07) | -0.37<br>(-0.41-<br>-0.33) | - | -0.14<br>(-0.18-<br>-0.09) | -0.32 (-0.36 -<br>-0.28) | 0.25<br>(0.21-0.30) | -0.30<br>(-0.34-<br>-0.26) |
| Relative<br>Humidity, % | 0.10<br>(0.06 -<br>0.15) | -0.52<br>(-0.55-<br>-0.48) | 0.04<br>(-0.00-0.09) | -0.14<br>(-0.18--0.09) | - | 0.31<br>(0.27-0.35) | -0.78<br>(-0.79-<br>-0.76) | 0.59<br>(0.56-0.62) |
| Precipitation,<br>mm | -0.03<br>(-0.07 -<br>0.02) | -0.01<br>(-0.05-0.04) | 0.25<br>(0.21-0.29) | -0.32 (-0.36<br>--0.28) | 0.31<br>(0.27-0.35) | - | -0.30<br>(-0.34-<br>-0.26) | 0.29<br>(0.24-0.33) |
| Sunlight<br>hours | -0.21<br>(-0.26 -<br>-0.17) | 0.50<br>(0.47-0.54) | -0.23<br>(-0.27-<br>-0.18) | 0.25<br>(0.21-0.30) | -0.78<br>(-0.79-<br>-0.76) | -0.30<br>(-0.34-<br>-0.26) | - | -0.80<br>(-0.81-<br>-0.78) |
| Cloud cover<br>(okta) | 0.1<br>(0.06 -<br>0.15) | -0.25<br>(-0.29-<br>-0.2) | 0.18<br>(0.14-0.23) | -0.30<br>(-0.34-<br>-0.26) | 0.59<br>(0.56-0.62) | 0.29<br>(0.24- 0.33) | -0.80<br>(-0.81-<br>-0.78) | - |

**Figure S3:**
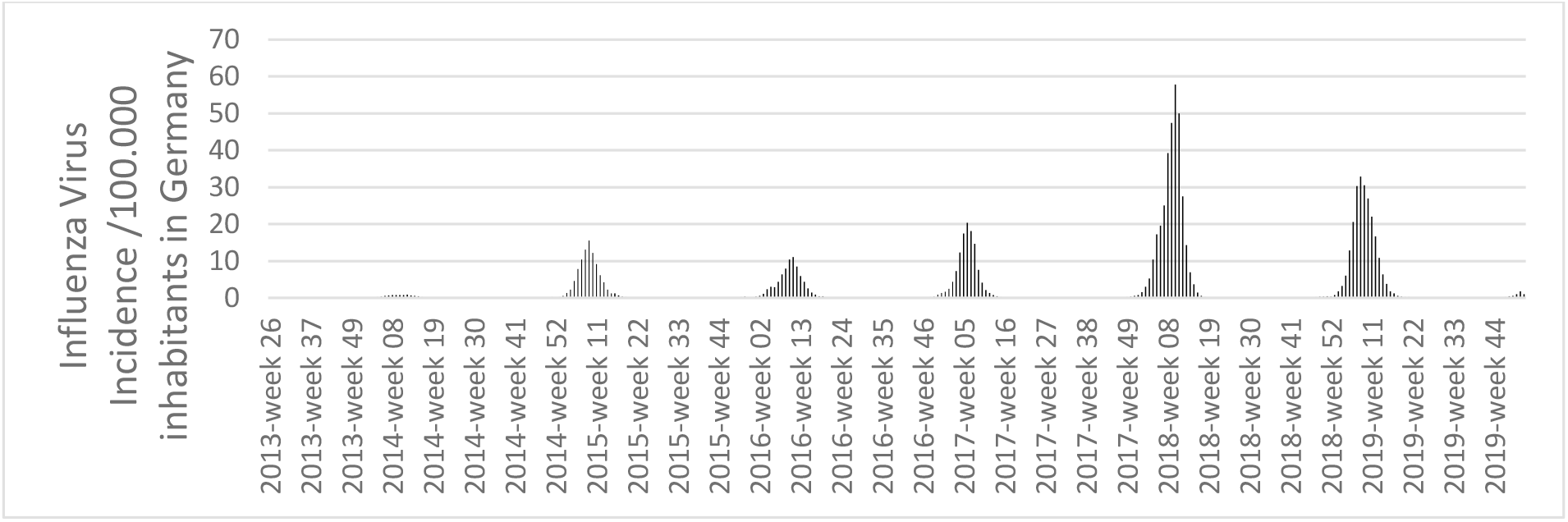
Influenza virus incidence per 100.000 inhabitants in Germany from June 2013 to December 2019. Data are publicly available from the Survstat@RKI 2.0 server. Robert Koch-Institut: SurvStat@RKI 2.0, https://survstat.rki.de, Query date: 22.06.2020.

